# Sidedness in unilateral orofacial clefts: A systematic scoping review

**DOI:** 10.1101/2023.09.06.23295169

**Authors:** Matthew Fell, Daniel Bradley, Ambika Chadha, Sophie Butterworth, Amy Davies, Craig Russell, Bruce Richard, Yvonne Wren, Sarah Lewis, David Chong

## Abstract

**Objectives:** An overview of the literature relating to the sidedness of unilateral cleft lip with or without cleft palate to map current knowledge on the cause and impact of directional asymmetry in the unilateral cleft lip phenotype.

**Data Sources:** Medline and Embase from inception to May 2023

**Study selection:** Studies including left and right unilateral cleft lip subgroups as a co-occurrence, an outcome or an exposure.

**Data analysis:** Narrative synthesis.

**Results:** Forty-one studies were eligible for inclusion; 12 studies reported cleft sidedness co-occurring with another phenotype, 12 studies report sidedness as an outcome and 17 studies as an exposure. Phenotypes which were reported to co-occur with either left or right sided clefts included congenital dental anomalies, handedness and additional congenital anomalies. Variables investigated as a potential cause of left or right sided clefts as an outcome included chromosomal anomalies, genetic variants and environmental factors. Outcomes investigated in relation to cleft sidedness as an exposure included facial anatomical features, facial growth, educational attainment, functional and psychological characteristics. More studies showed worse outcomes in right sided clefts versus left sided clefts than vice versa, although studies were inconsistent, and a quality assessment was not performed.

**Conclusions:** The field of cleft sidedness research is expanding and there are promising early findings to differentiate cause and outcome by sidedness of the cleft.

## INTRODUCTION

The prevalence of left sided unilateral cleft lip with or without cleft palate (UCL/P) is consistently observed to be twice that of right sided UCL/P across multiple studies and populations ^1^.

Lateralization to left or right is one of the three major axes of polarisation during embryological development, together with anterior/posterior and dorsal/ventral orientations. The developmental timing of lateralization in the human body occurs after the other two axes of polarization have occurred, shortly after gastrulation at the fourth week of gestation ^2^. Whilst most aspects of the body are symmetrical about the sagittal plane, there is asymmetry of the organs in the thoracic and abdominal regions and hemispheric specialisation in the brain ^3^. The asymmetry of body organs is driven by a complex cascade of molecular pathways including Nodal and the transcription factor Pitx2 ^4^. The biological mechanisms that determine the laterality axis in embryological development are thought to involve the rotation of motile cilia which create a leftward flow in the node (a midline pit containing cilia) during gastrulation ^5^.

Clefts of the embryological primary palate most commonly occur as a left unilateral cleft lip, a right unilateral cleft lip or a bilateral cleft lip in a ratio of 6:3:1^6^. The embryological primary palate (upper lip and alveolus up to the incisive foramen) is formed from the fusion of medial nasal and maxillary processes between 4-7 weeks of gestation, which coincides with the timing of organogenesis ^7^. This process is dependent upon the migration of cells from neural crest origin, followed by correctly timed growth to form lateral swellings and finally a complex series of fusions ^8^. A unilateral cleft of the lip is characterised by a gap in the upper lip through the left or right philtrum to the underlying alveolus, between the ipsilateral lateral incisor and canine tooth, following the line of suture incisiva up to the foramen incisivum.

The embryological secondary palate (hard and soft palate posterior to the incisive foramen) develops later between 8-12 weeks of gestation, from paired outgrowths on the maxillary processes called the palatal shelves. The palatal shelves are initially positioned vertically and subsequently reorientate into the horizontal plane prior to midline fusion, a process which is often asynchronous with one shelf reorientating before the other ^9^. An isolated complete cleft of the secondary palate occurs in the midline and is characterised by the vomerine portion of the nasal septum being unfused to either palatal shelves (as is also the case in a bilateral cleft lip and palate). A unilateral cleft of the lip and palate is characterised by a lateralized cleft on either the left or right within the hard palate, with fusion of the vomer to the non-cleft palatal shelf, whereas the cleft of the soft palate remains in the midline with the musculature of the velum and uvula failing to unite. This phenomenon is illustrated in the LAHSHAL classification^10^ where clefts of the lip, alveolus and hard palate are coded twice to enable documentation of involvement of a left or a right side, compared to a cleft of the soft palate which is coded only once due to its occurrence in the midline.

Our aim was to gain an understanding of the current knowledge base on the cause of directional asymmetry in UCL/P and its impact on outcomes. Scoping reviews are characterized by a systematic approach to map evidence and are well suited to clarifying key concepts, summarizing bodies of heterogenous literature, identifying research gaps and making recommendations for further research ^11^. The key difference from a systematic review is that a scoping review provides an overview of the literature regardless of quality, therefore a quantitative synthesis or meta-analysis is not undertaken ^12^.

Our objectives in this scoping review were threefold: firstly, to systematically search the published literature for studies which have reported on sidedness in UCL/P, secondly to assess what is known about the determinants of sidedness in UCL/P via embryological, genetic or environmental processes and thirdly to determine whether there are any differences in outcomes reported when stratified by sidedness of UCL/P.

## METHODS

### Protocol and registration

This study followed the Joanna Briggs Institute guidance for conducting a systematic scoping review ^11^ and was reported according to PRISMA-ScR guidance ^12^ (see supplementary Table 1). A full protocol of this review is available from the PROSPERO systematic review register (registration number CRD42022333191: https://www.crd.york.ac.uk/prospero/display_record.php?RecordID=333191).

### Identification of studies

Eligible studies were defined as full-text publications reporting on humans or animals born with UCL/P. The protocol included descriptive and analytical study designs and both primary and secondary data. We included case series and case reports, but commentaries or editorials were not included (see Supplementary Table 2 for inclusion and exclusion criteria).

The variable of interest was unilateral cleft sidedness, defined as a left or right sided UCL/P. We focused on the sidedness of unilateral cleft lip, rather than the sidedness of clefts in the alveolus or hard palate because of the external nature of the lip, making it easier and more reliable to determine left versus right. Studies were categorized depending on whether they considered UCL/P sidedness as a co-occurrence, an outcome or an exposure. Where UCL/P sidedness was a co-occurrence, studies reported the prevalence of additional phenotypes. Where UCL/P sidedness was an outcome, studies reported variables involved in the aetiology of a cleft. Where UCL/P sidedness was an exposure, studies reported anatomical and functional features.

The databases searched included Medline and Embase from inception to 22^nd^ May 2023. The search was tailored individually to each database with input from a University Librarian (see Supplementary Table 3 for search strategies) and there were no language restrictions. The search focused on published literature and did not include grey literature. In addition, manual searches of reference lists of all studies included in the review were performed.

Titles and abstracts were reviewed independently in the first round of screening by two reviewers (MF/DB) according to the specified inclusion and exclusion criteria. Differences were resolved through discussion to reach a consensus. In the second round of screening, full text screening was performed independently by the same two reviewers for inclusion and any disagreements resolved through discussion. When multiple reports of a single study were identified, the report with the greatest number of patients was selected. The Rayyan web application was used to facilitate the screening process ^13^.

### Data extraction and synthesis

Data was extracted via Microsoft Forms into an Excel spreadsheet. Data extracted included: title, authors, publication year, country of study population, study design, sample size, confounding factors and reported results. Due to the potential for studies to report multiple outcomes and exposures, we reported all pertaining to UCL/P sidedness for transparency, to avoid selecting only positive results. Due to the heterogeneity of study designs it was not possible to perform a meta-analysis, so results were reported in tables. A descriptive summary and narrative synthesis of the included studies was performed in accordance with published guidance ^14^.

## RESULTS

### Study Selection and Study Characteristics

A flowchart for the article review process is shown in Figure 1. A total of 2790 citation records were identified from searching the two databases and a manual search of included articles identified 11 additional studies. After exclusions, 41 studies were included in the scoping review. The earliest study to be included in the review was published in 1985 ^15^. Forty studies were published in the English language and one study was published in German ^16^. Forty studies involved human subjects and one study used a murine model ^17^. Participants in the studies were reported to have the following unilateral cleft lip sub phenotypes: unilateral cleft lip with or without cleft palate (UCL/P: n=17), unilateral cleft lip and palate (UCLP: n=14), unilateral cleft lip with or without a cleft of the alveolus (UCL+-A n=4) and finally distinct populations of both UCLP and UCL+-A (n=16).

**Figure 1:**
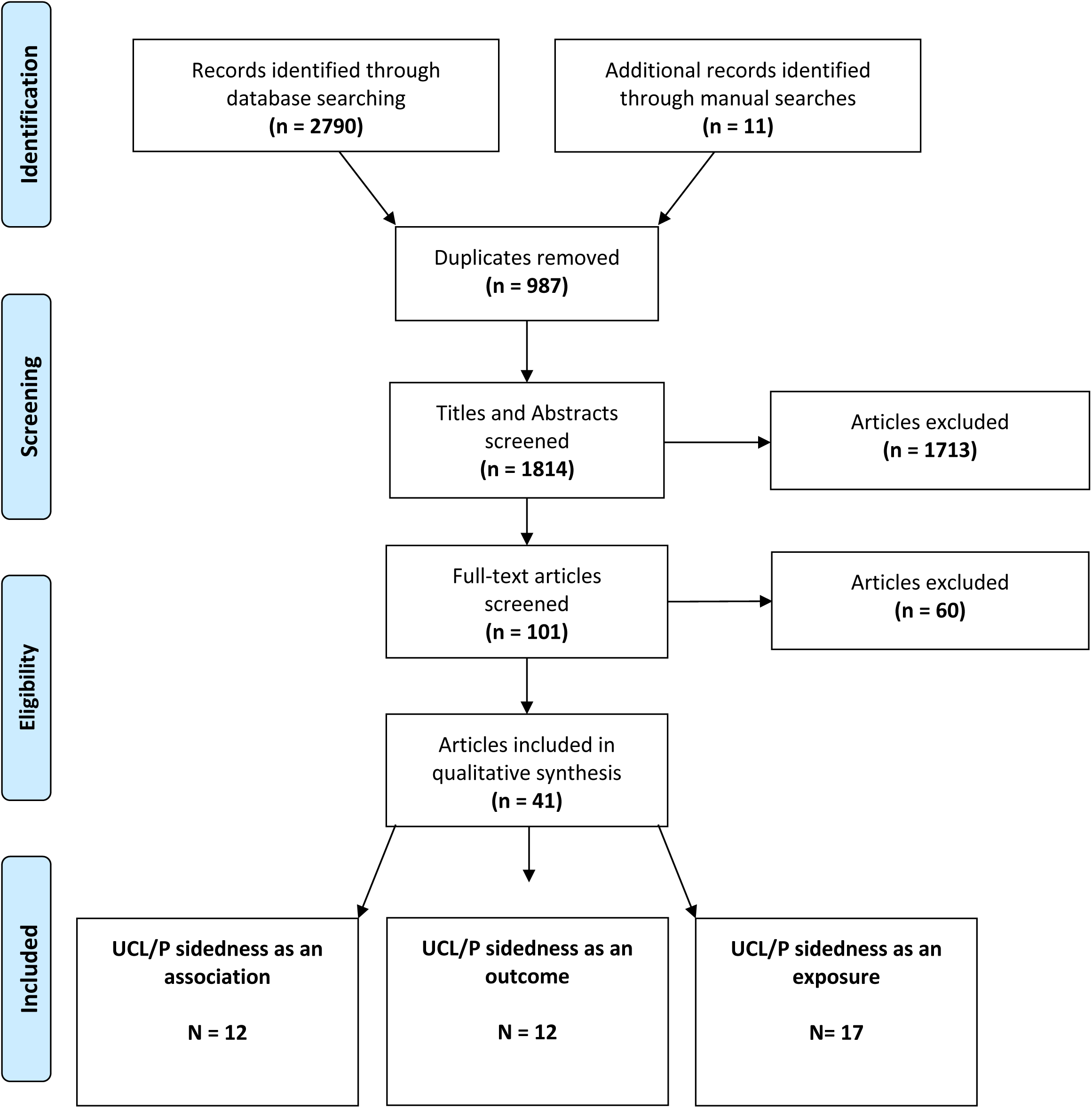
A flow chart of the search strategy and study selection.

Twelve studies reported UCL/P sidedness as a co-occurrence (Table 1), 12 studies reported UCL/P sidedness as an outcome (Table 2) and 17 studies reported UCL/P sidedness as an exposure (Table 3). In total, 9,104 left sided UCL/P (66%) and 4,890 right sided UCL/P (34%) were reported from the 41 studies, and whilst we cannot exclude patient overlap, this does align with the anticipated predominance of left sided UCL/P.

**Table 1:**
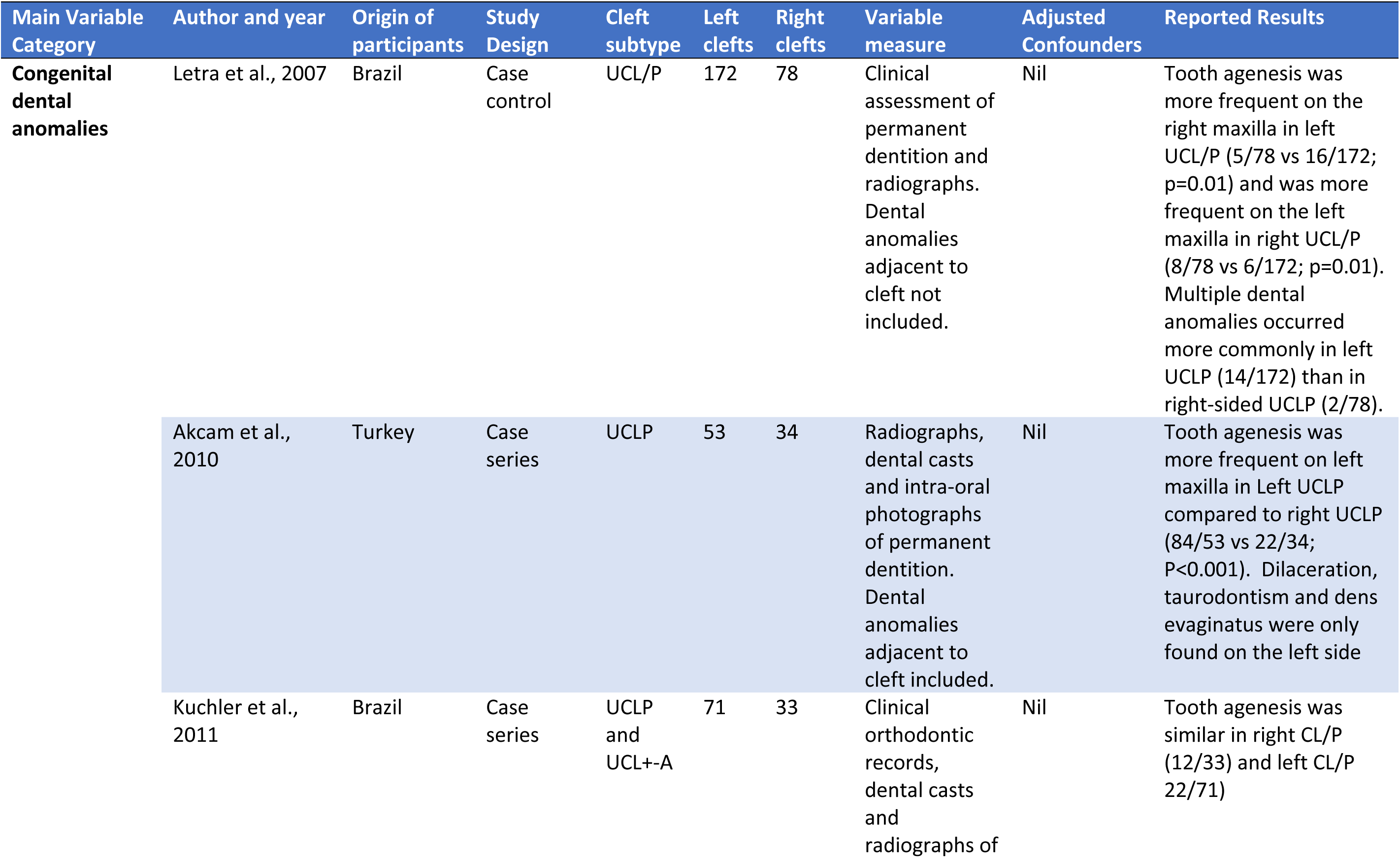
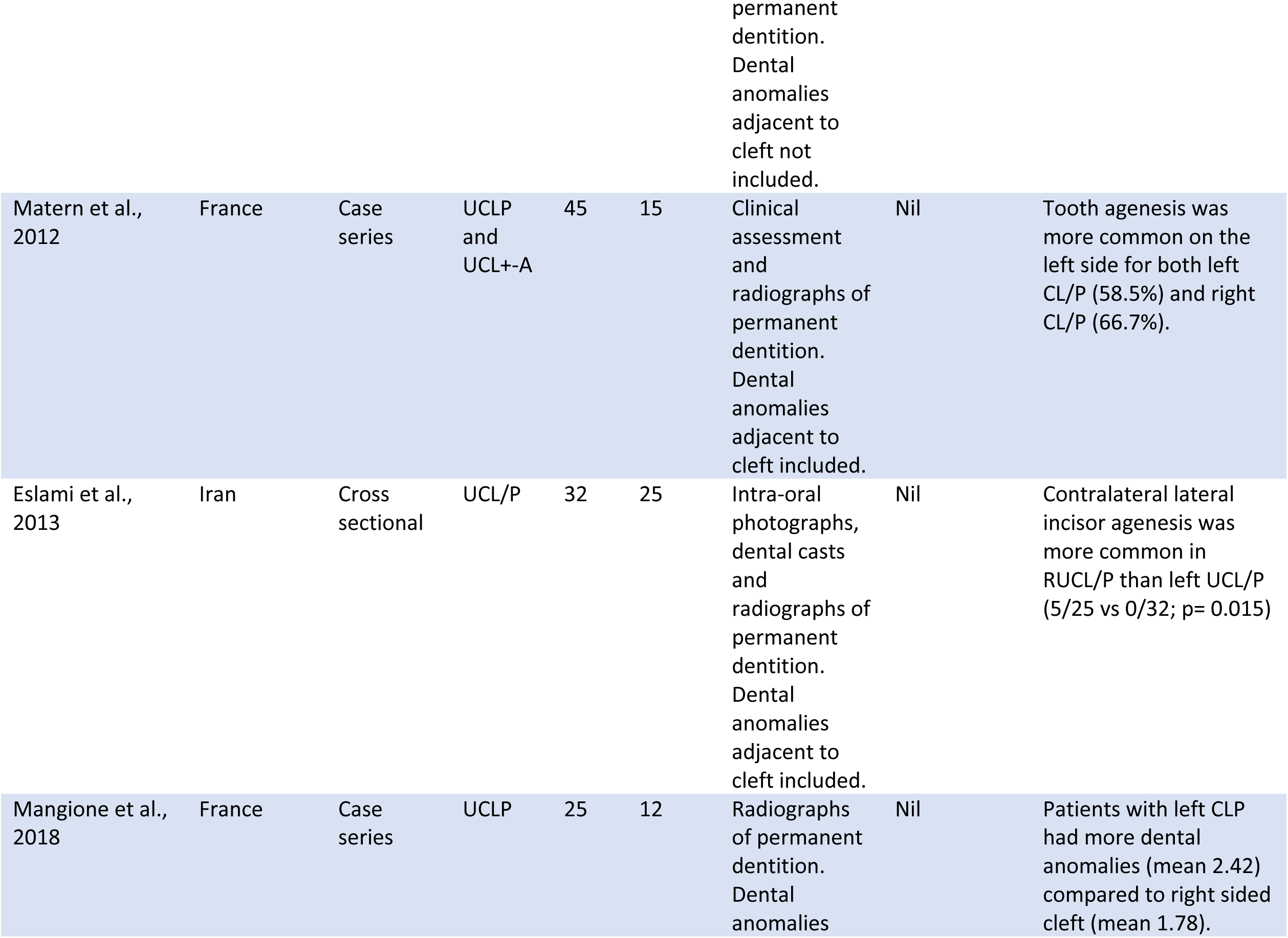
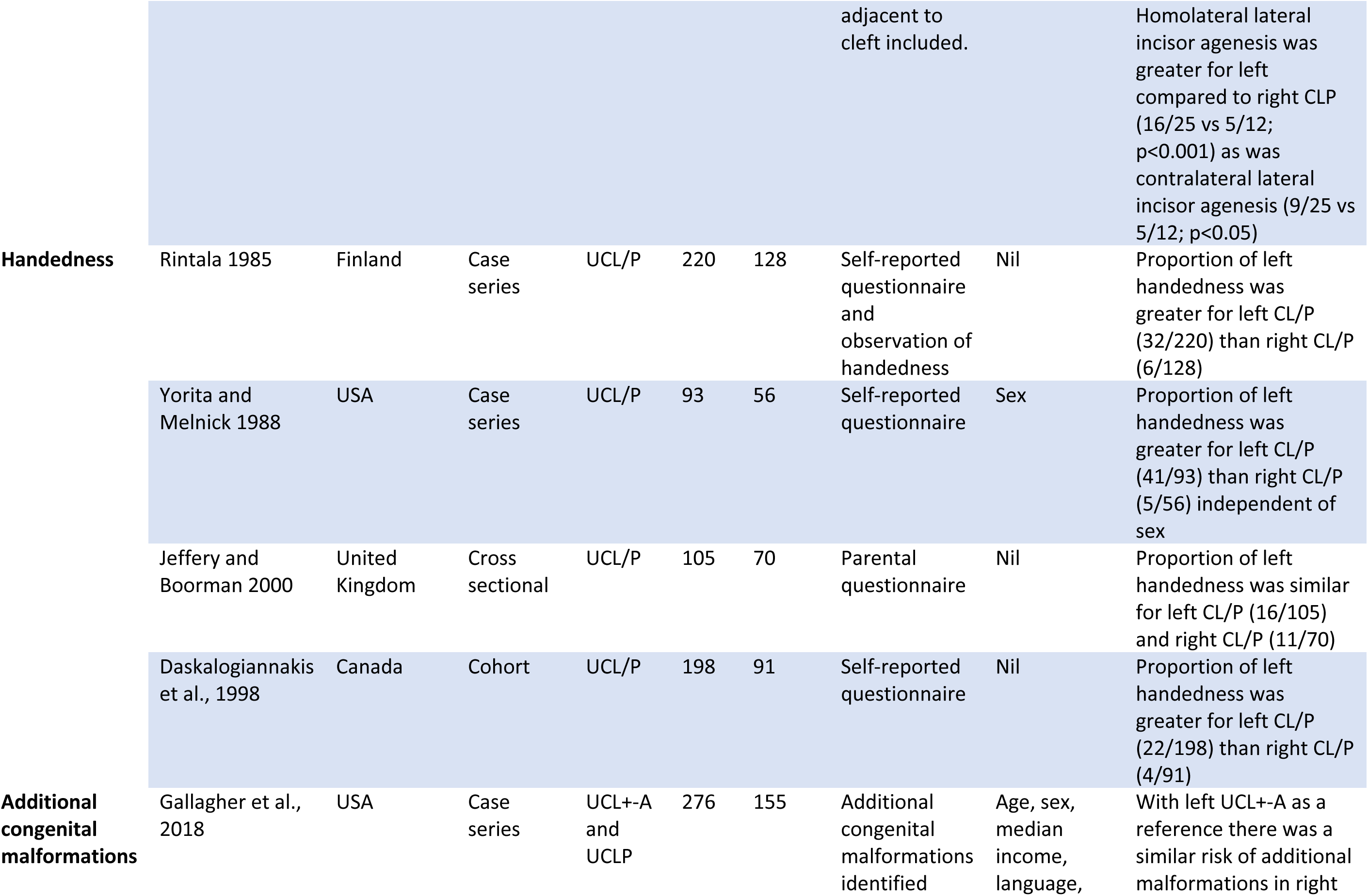
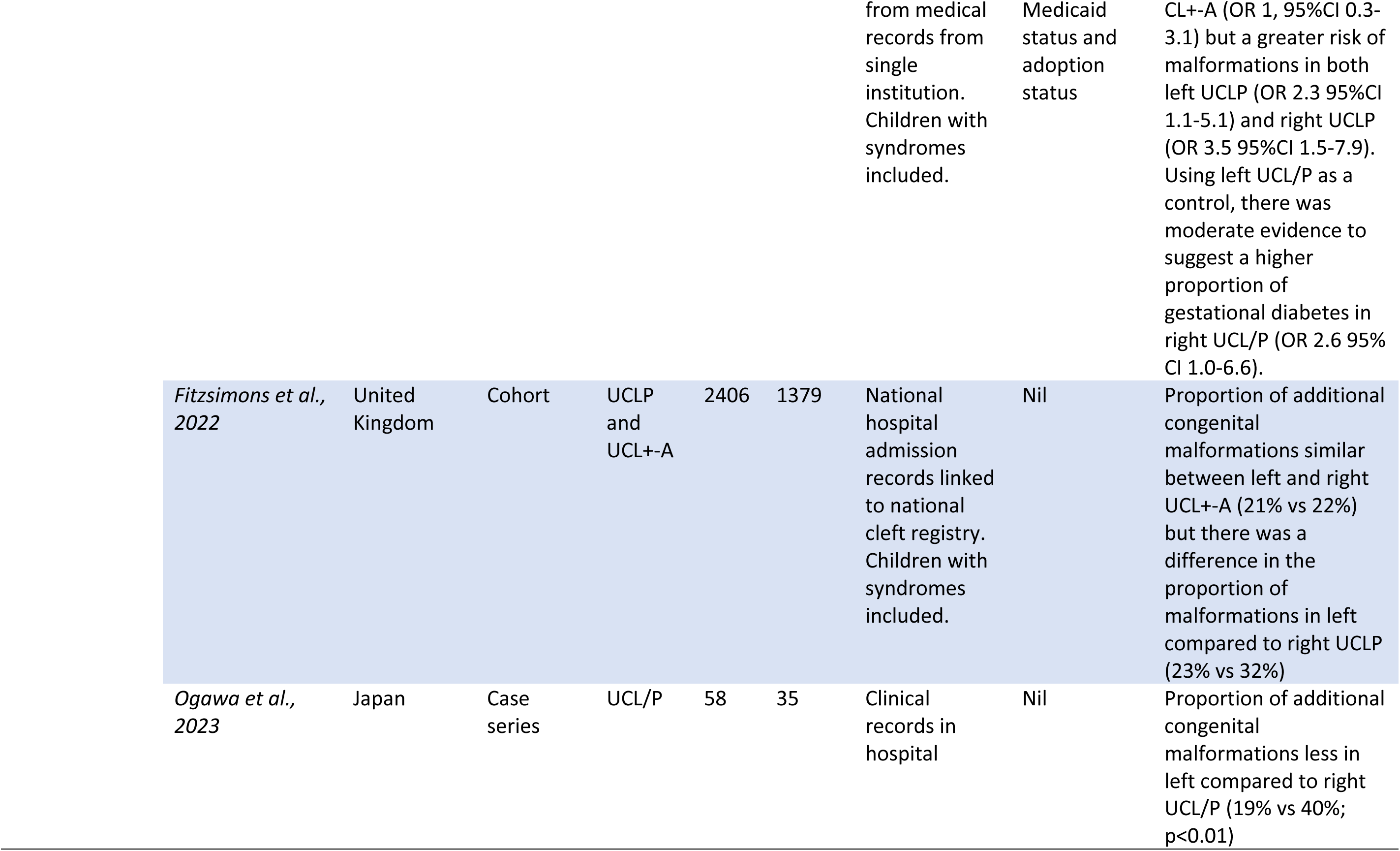
Included studies reporting left versus right sided cleft as a co-occurrence.

**Table 2:**
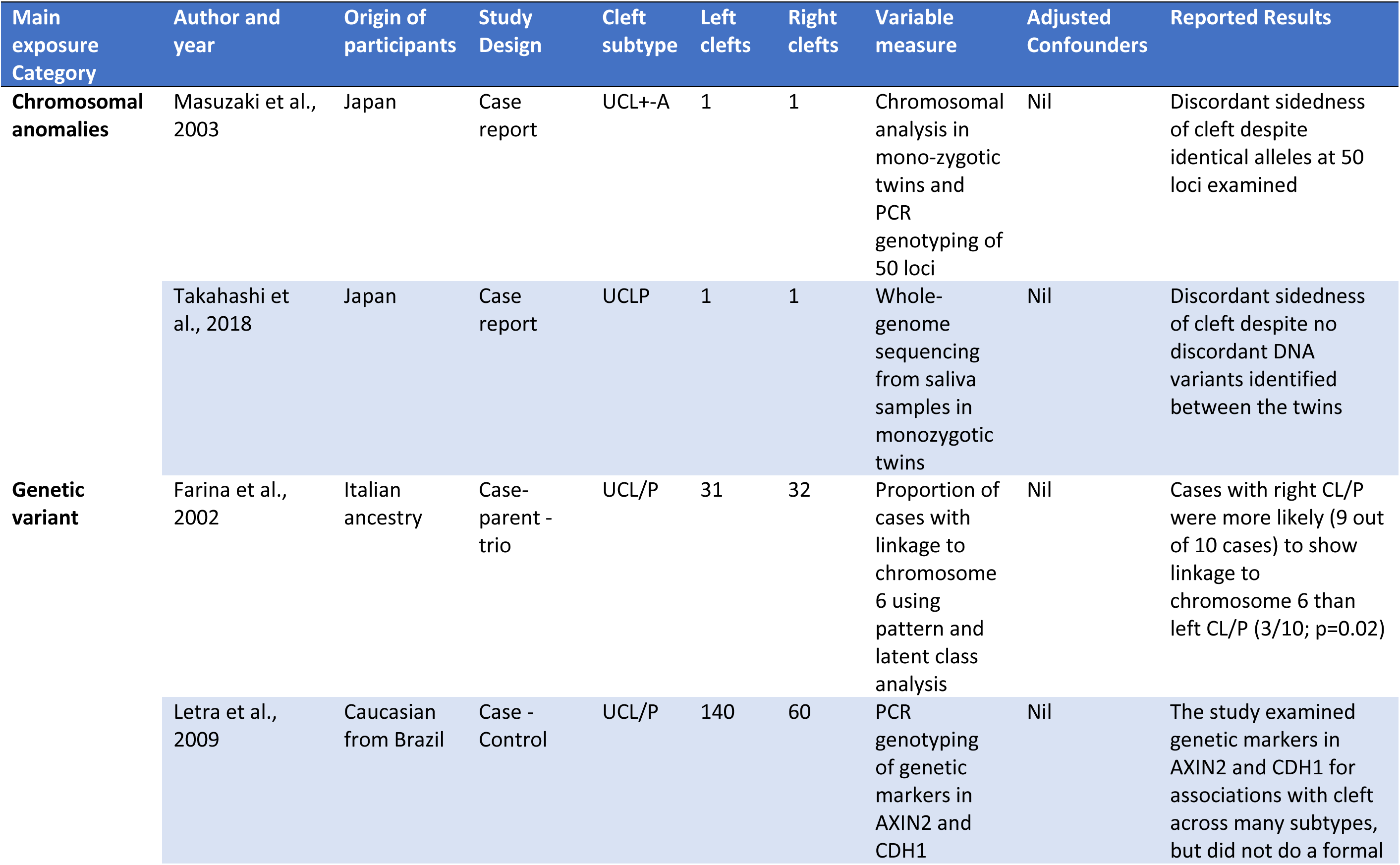
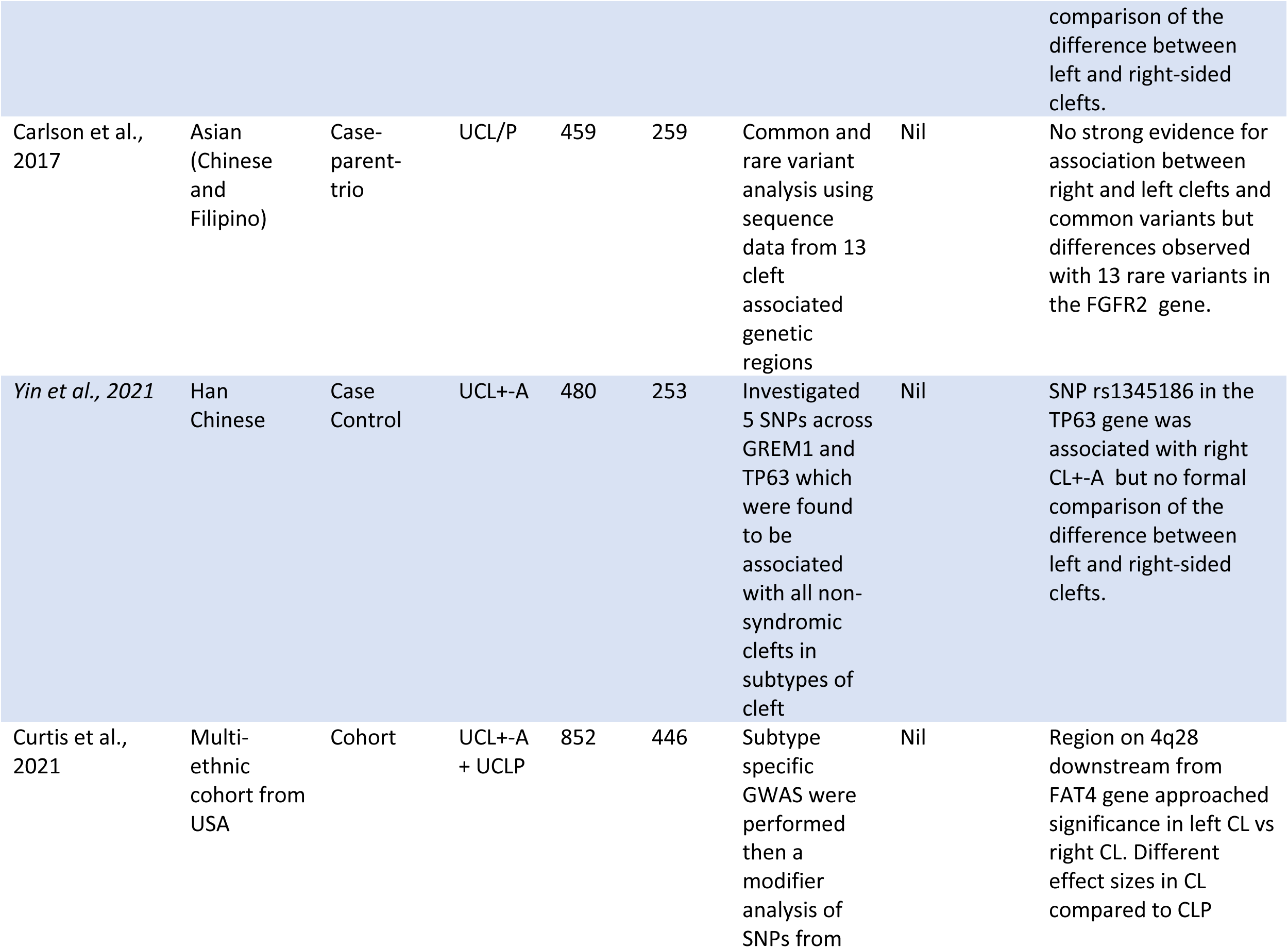
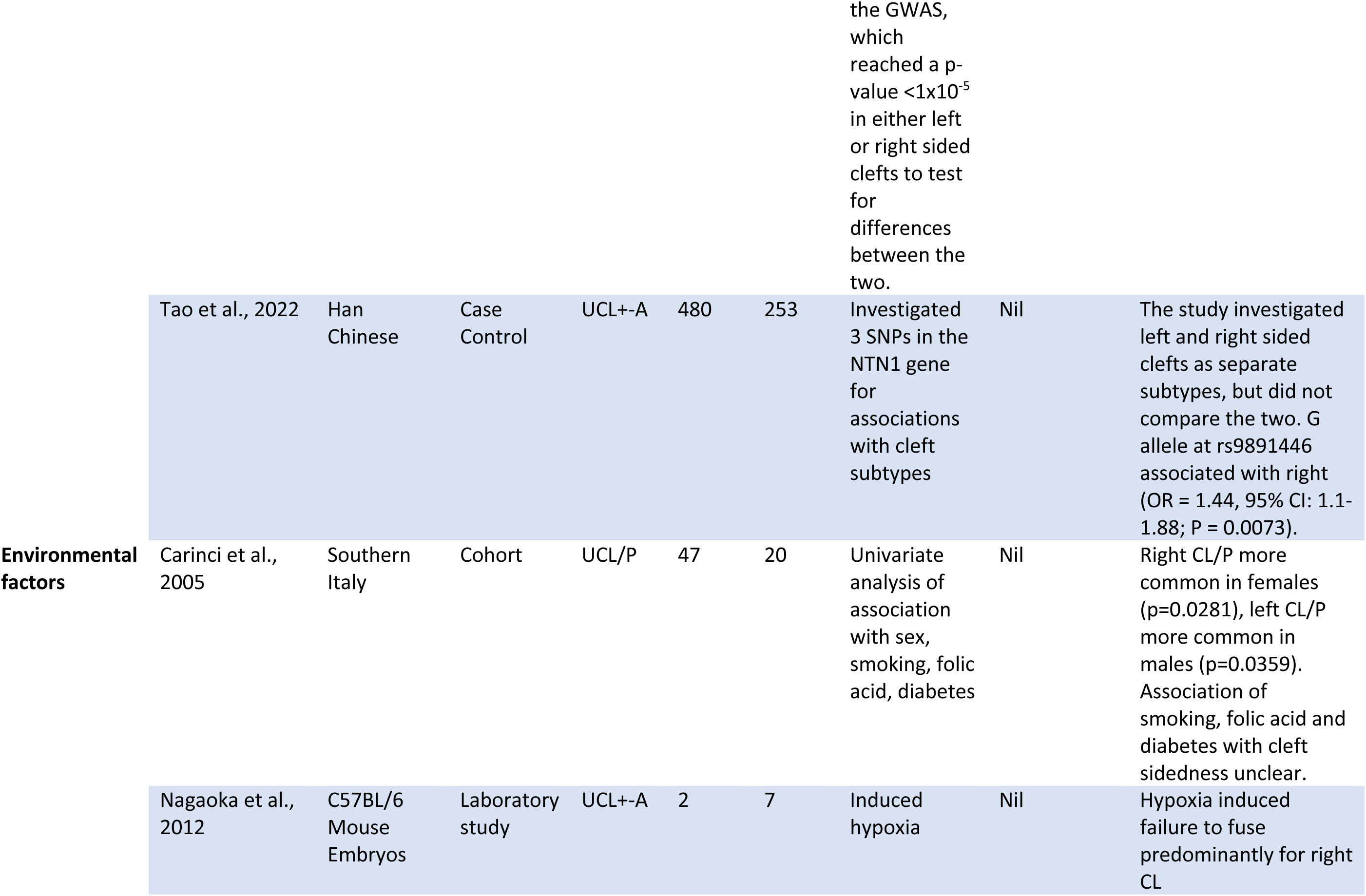
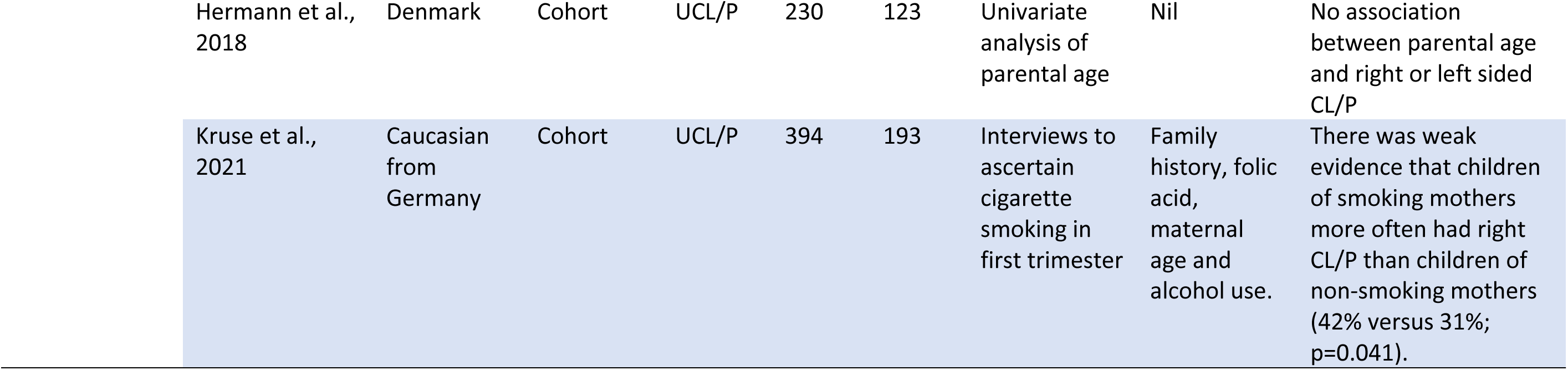
Included studies reporting left versus right cleft as an outcome.

**Table 3:**
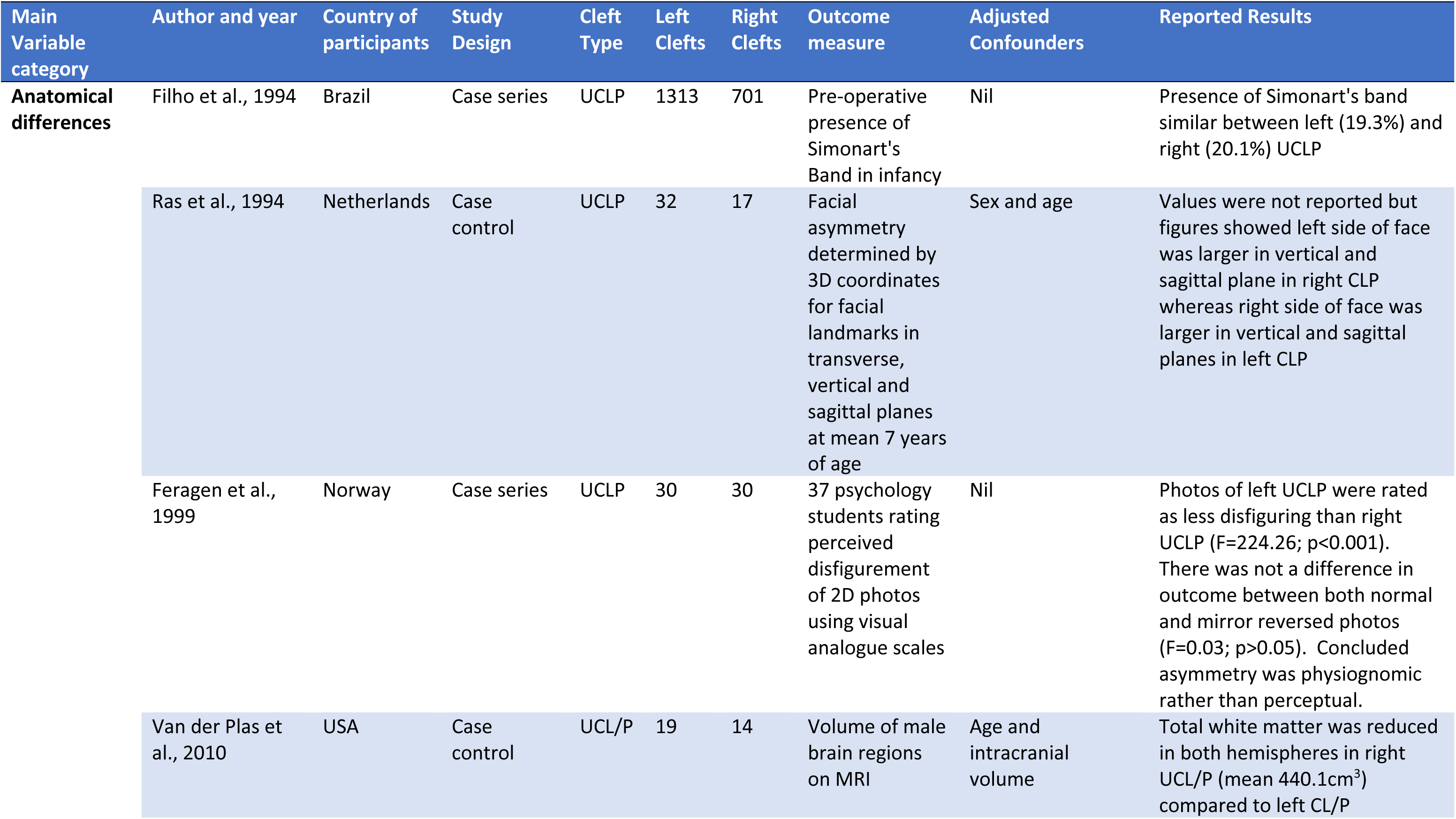
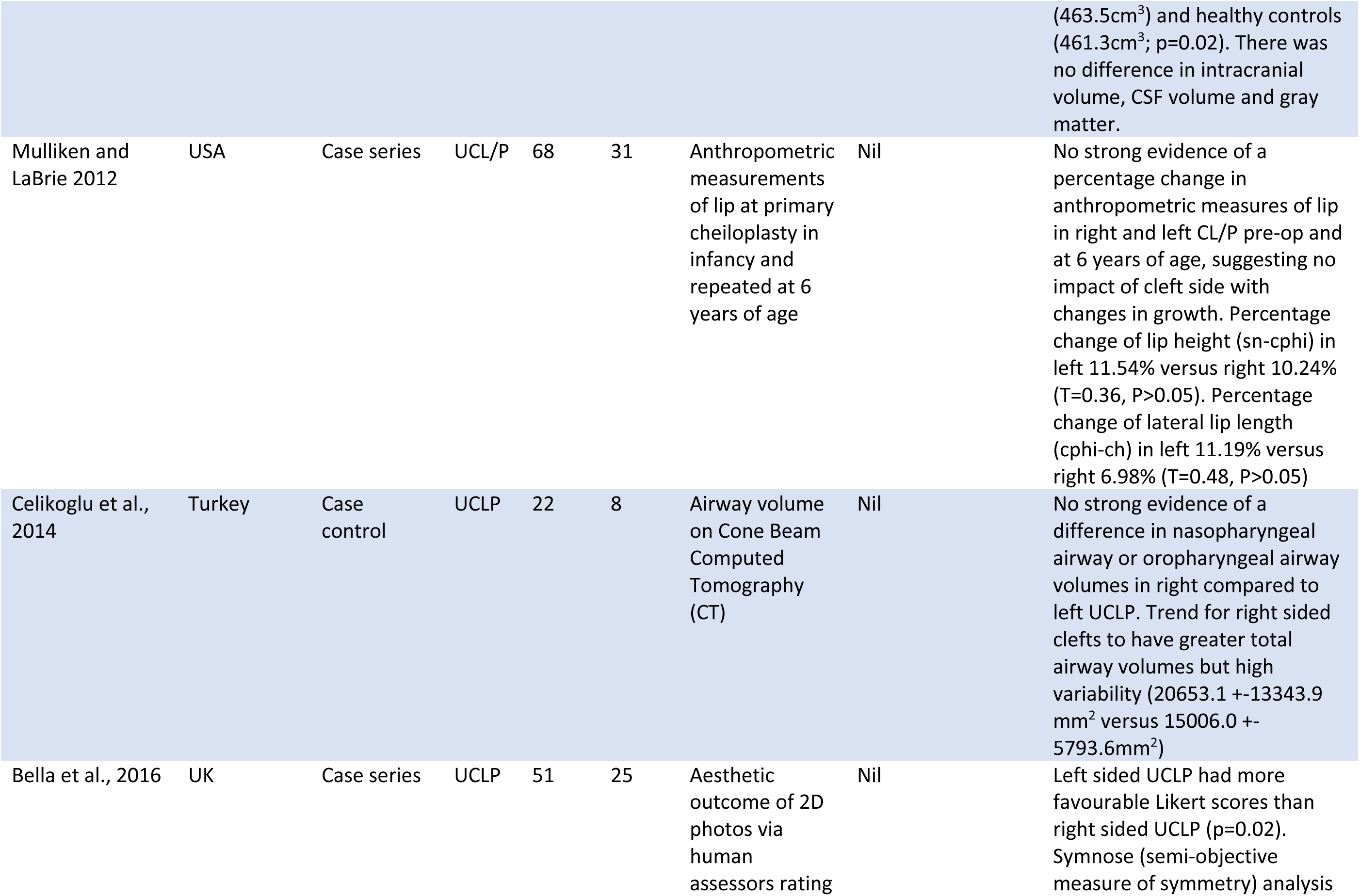
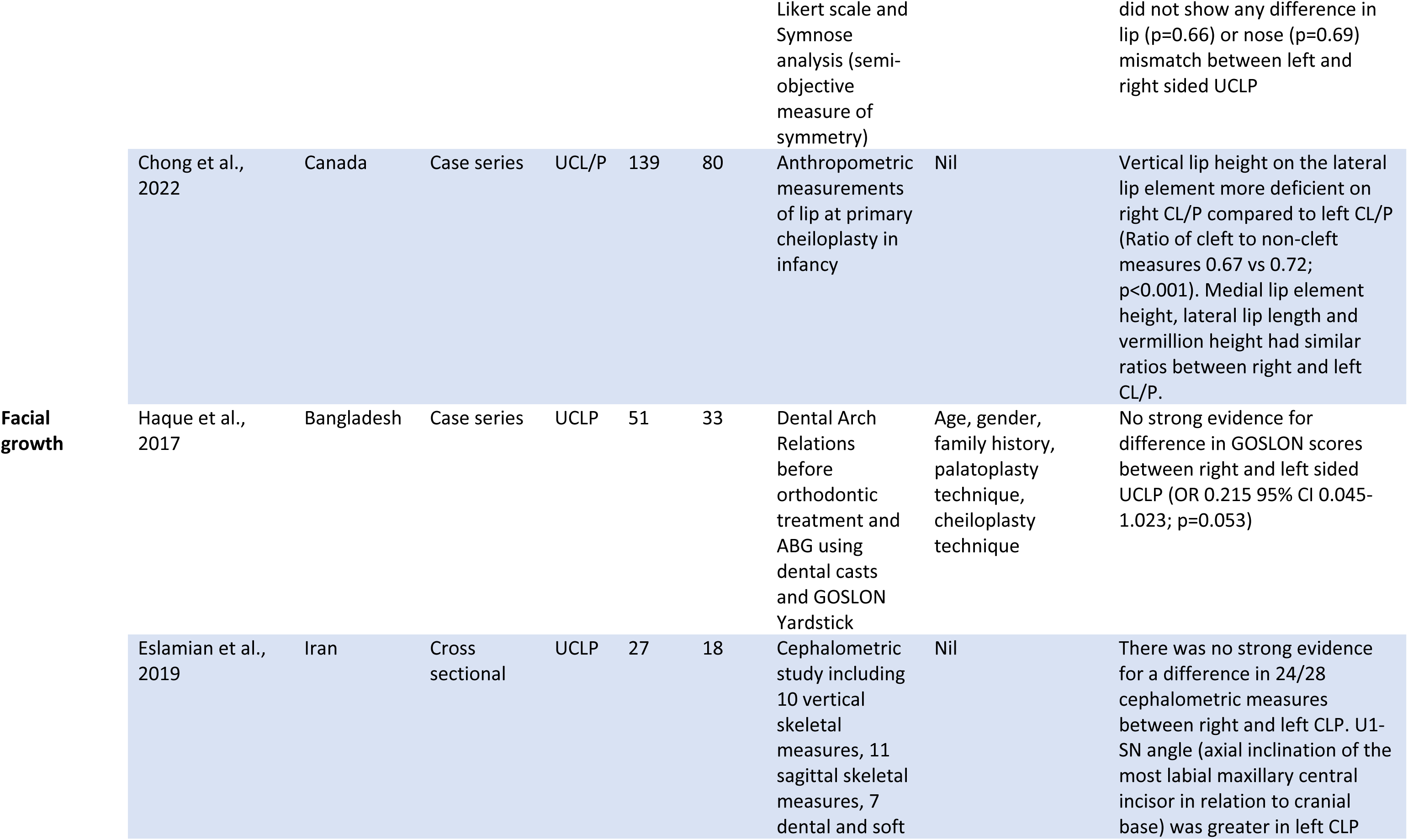
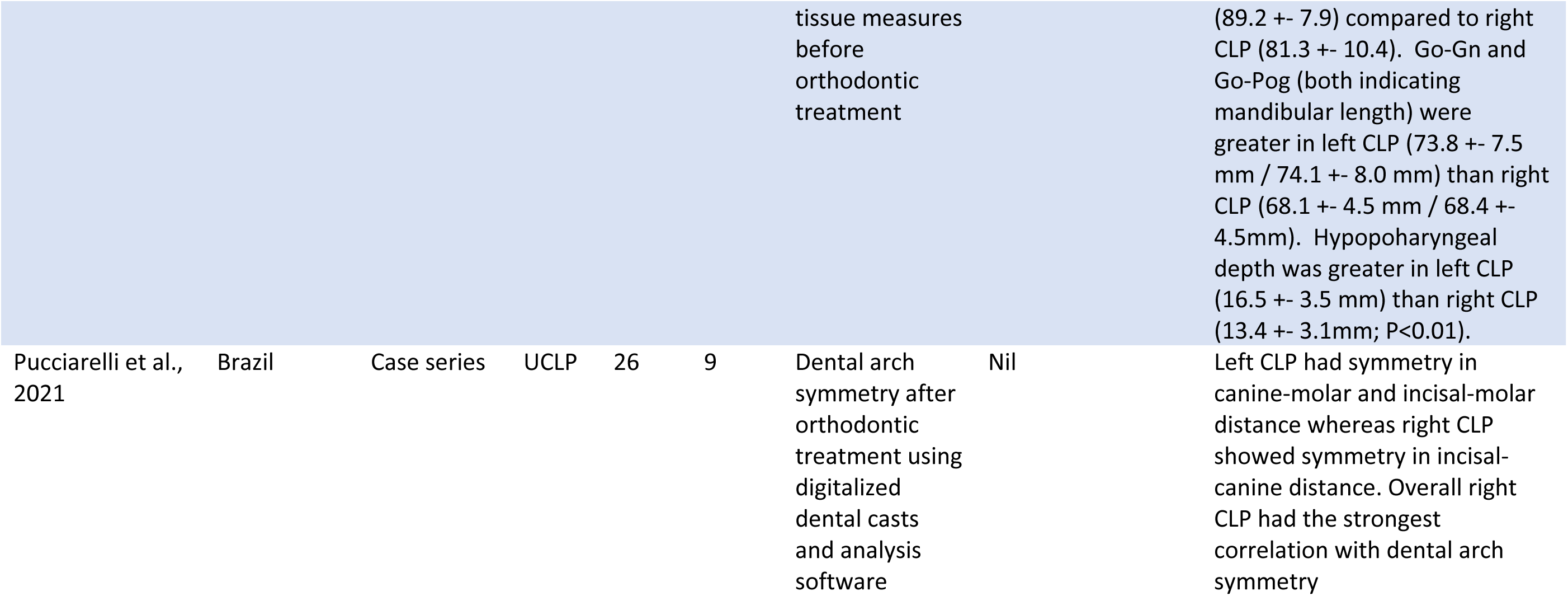
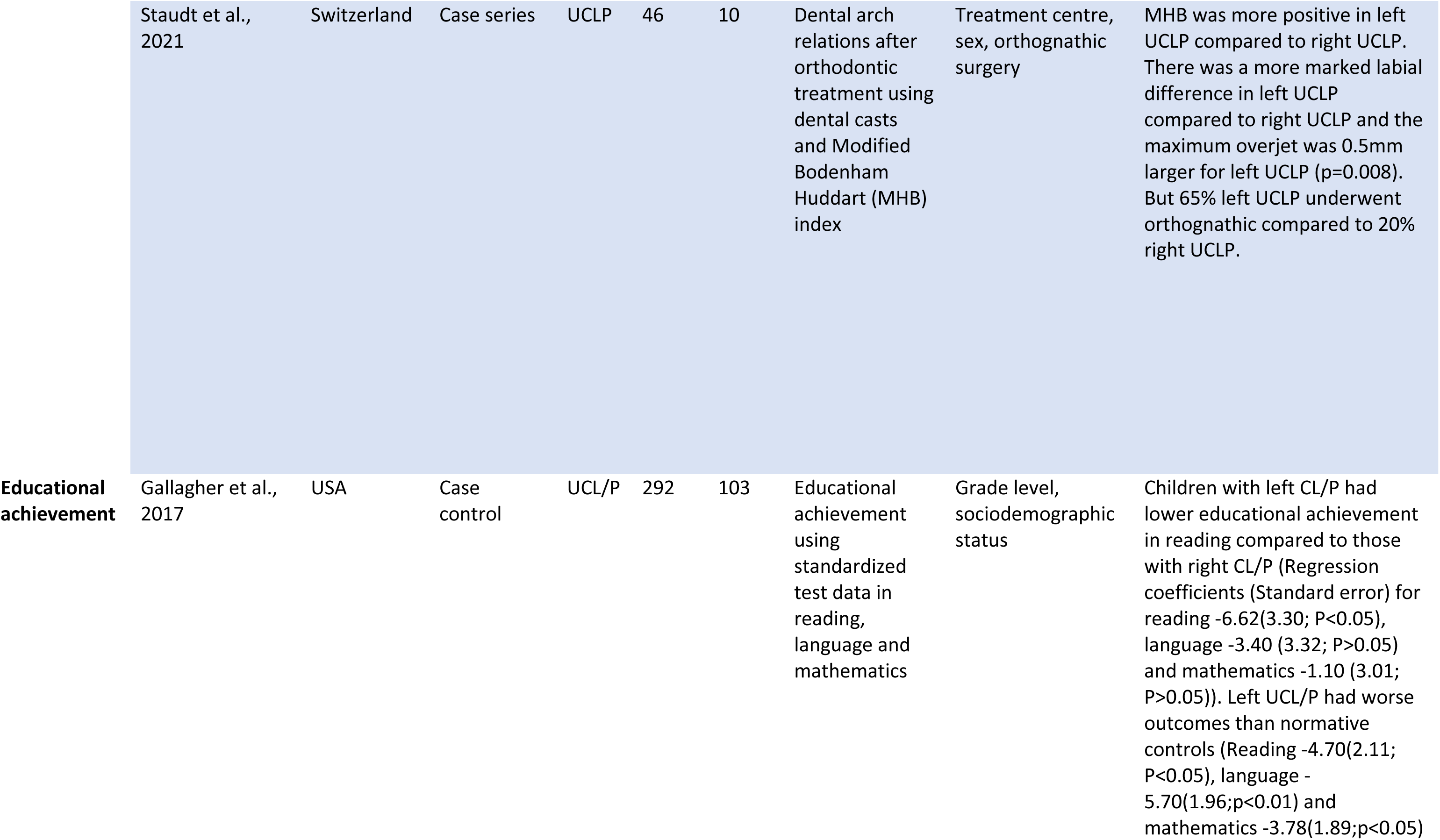
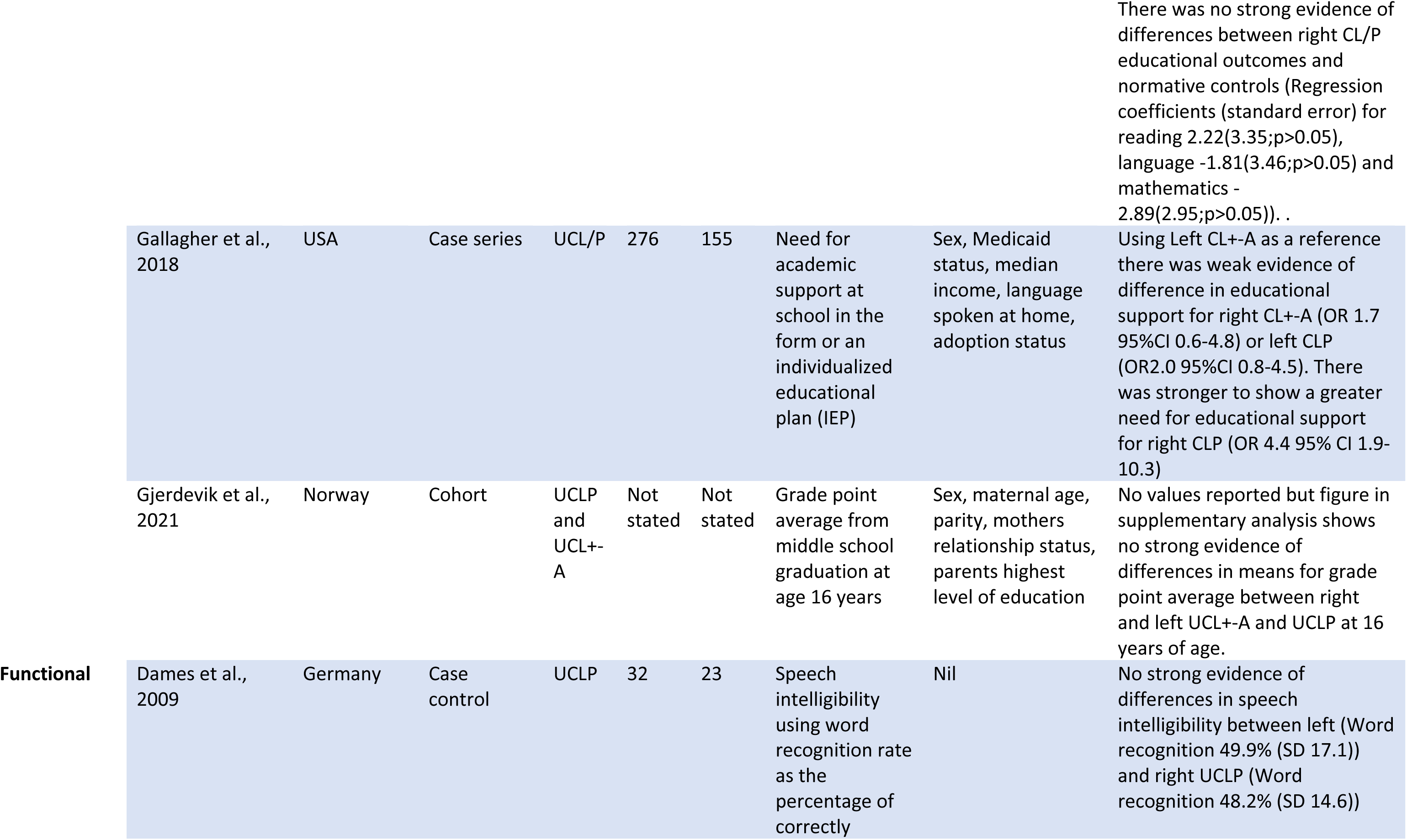
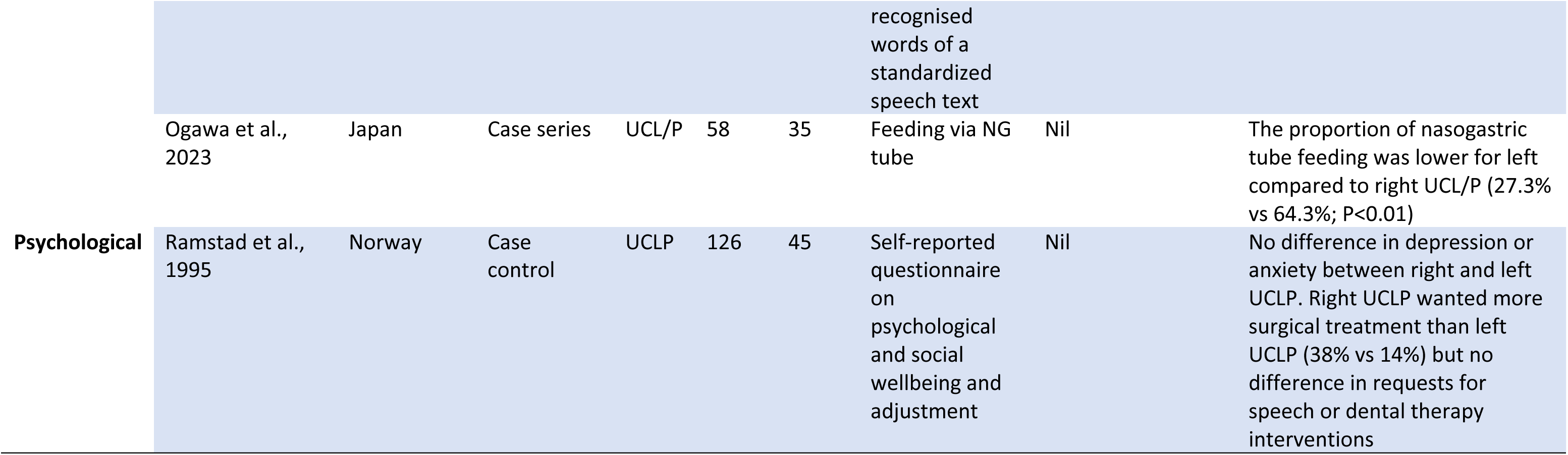
Included studies reporting left versus right sided cleft as an exposure.

### UCL/P sidedness as a co-occurrence

Twelve studies reporting other phenotypes which co-occurred with either left or right sided UCL/P were published from 1985-2022 (Table 1). Study designs included case series (n=7), cohort (n=2), case-control (n=1) and cross-sectional studies (n=2). Phenotypes investigated as co-occurring with sidedness were categorized as congenital dental anomalies (n=6), handedness (n=4) and additional congenital anomalies (n=2).

There were six studies reporting the presence of congenital anomalies of permanent dentition. The studies were heterogenous in terms of dental anomaly classification and inclusion of dental anomalies adjacent to the cleft in the analysis. Tooth agenesis was the anomaly with the highest frequency and was reported in all six studies. A higher frequency of tooth agenesis in left compared to right UCLP was reported in three studies ^18–20^ whereas one study found no difference in tooth agenesis between left or right UCLP ^21^. Two studies reported right sided UCL/P being more likely to have contralateral incisor agenesis^22,23^ and Matern et al (2012) proposed a theory that this could be due to right sided UCL/P being a lesser form of bilateral BCL/P.

Four studies reported on the co-occurrence of handedness with UCL/P sidedness. The proportion of left handedness was greater in left UCL/P compared to right UCL/P in three studies ^15,24,25^ whereas another study found no difference in handedness between left or right UCL/P ^26^.

Additional congenital anomalies were found to be less prevalent for left compared to right UCL/P in the three studies publishing on this variable ^27–29^. When Fitzsimons et al. and Gallagher et al stratified by cleft subtypes, ^27,28^ they found this trend was true only for UCLP and not for UCL+-A.

### UCL/P sidedness as an outcome

Twelve studies reported UCL/P sidedness as an outcome and were published from 2002-2022 (Table 2). Study designs included cohort (n=4), case-control (n=3), case-parent-trios (n=2), case reports (n=2) and a single laboratory study (n=1). Variables investigated were categorized as genetic variants (n=6), chromosomal anomalies (n=2) and environmental factors (n=4).

Two case reports from Japan performed chromosomal analysis in pairs of monozygotic twins born with discordant sides of UCL/P ^30,31^. Masuzaki et al reported identical alleles across the 50 sites investigated in the twins and Takahashi et al, reported no differences between the twins following whole genome sequencing despite the different UCLP sidedness profile.

Four candidate gene studies investigated whether specific genetic loci were associated with left and right-sided UCL/P subgroups. These loci investigated were chromosome 6 ^32^, TP63 gene on chromosome 3 ^33^, CDH1 gene on chromosome 16 ^34^ and NTN1 gene on chromosome 17 ^35^. Only the study by Farina et al. ^32^ of chromosome 6 compared left with right sided UCL/P, whereas the other studies reported comparisons of sidedness subgroups with controls. Carlson et al. ^36^ investigated common and rare variants, using sequencing data from 13 cleft associated regions, to determine whether they influenced left versus right sided UCL/P; they found evidence for 13 rare variants in the FGFR2 gene. Curtis et al., ^37^ conducted subtype specific GWAS then ran a modifier analysis of SNPs from the GWAS, which reached a p-value <1×10-5 in either left or right sided UCL/P to test for differences between the two. They reported that a region downstream of the FAT4 gene on chromosome 4 approached significance in an analysis of left versus right sided UCL/P, raising the potential for distinct genetic modifiers for sidedness.

Environmental factors influencing cleft laterality were investigated in four studies. Maternal cigarette smoking was associated with more right sided UCL/P compared to left sided UCL/P in a 2021 German cohort ^38^, whereas the relationship of smoking to sidedness was unclear in a smaller Italian cohort ^39^. Hermann et al. ^40^ reported no association with parental age and UCL/P sidedness. A study using induced hypoxia in a mouse model reported a failure of fusion predominantly resulting in right sided UCL+-A ^17^.

### UCL/P sidedness as an exposure

Sixteen studies reported UCL/P sidedness as an exposure by investigating the impact of sidedness on cleft related outcomes (Table 3). Included studies were published over three decades from 1994-2023. Study designs were all based on human participants and included case series (n=8), case-control (n=6), cohort (n=1) and cross-sectional (n=1). Outcomes investigated were categorized as anatomical differences (n=7), facial growth (n=4), educational attainment (n=2), functional (n=2) and psychological (n=1).

Studies reporting anatomical differences according to UCL/P sidedness covered a broad variety of outcome measurement including anthropometric measurements of the face, perceptive facial attractiveness, brain volumes and airway volumes. Four studies reported anatomical differences of the lip with right sided UCL/P more often associated with; hypoplasia of the lateral lip element ^41^, more pronounced disfigurement ^42,43^ and asymmetry ^44^ compared to left sided UCL/P. Ras et al ^44^ found the non-cleft side of the face to be asymmetrically larger than the cleft side of the face in the vertical and sagittal planes in both left and right sided UCLP. Two studies of facial anatomy reported no differences between right and left sided UCL/P ^45,46^. It is worth noting that the anthropometric measurements were not consistent across studies, therefore direct comparison across studies is not possible. White matter brain volume was reported to be lower for males with right UCL/P compared to males with left UCL/P ^47^. Volumetric analysis of airway anatomy reported no difference between left and right sided UCLP.

Facial growth was measured using cephalometric measurements, arch symmetry assessment and dental arch relationship analysis. A study investigating cephalometric measurement of facial growth reported no difference in most of the 28 measurements reported between left and right UCLP but within the four measurements noted to be different there was greater mandibular length in left sided compared to right sided UCLP ^48^. Right UCLP was noted to have a stronger correlation with dental arch symmetry in a small Brazilian series ^49^. Dental arch relationships with sidedness of cleft were inconclusive; in the study by Staudt et al. ^50^ greater maxillary growth for left sided UCLP was reported using Modified Huddart Bodenham scores and overjet whereas Haque et al. ^51^ reported no difference in GOSLON scores between left or right sided UCLP.

Educational attainment in reading was reported to be lower for left versus right sided UCL/P in a case control study performed in the USA ^52^. However, contradictory findings by the same author in a later paper, showed a greater need for academic support in school in right compared to left sided UCL/P involving a different study population (Gallagher et al. 2018). One hypothesis provided by the authors for the difference between the studies was that their first study excluded patients who had additional congenital anomalies. No difference in grade point averages at 16 years of age were noted among those with left versus right sided UCL/P in a Norwegian cohort study ^53^.

A study published in German, which investigated functional outcomes including speech, reported no difference in intelligibility between left and right UCLP ^16^. Another study conducted in Japan investigated feeding and reported a lower proportion of tube feeding required for infants born with a left versus right sided UCL/P ^29^. A single study investigating psychological adjustment reported people born with right sided UCLP being more likely to request further surgical intervention compared to left sided UCLP ^54^.

## DISCUSSION

### Summary of evidence

The phenomenon of directional asymmetry in UCL/P with a 2:1 left to right ratio is universally accepted and established, yet despite this there has been relatively little research in the field of UCL/P sidedness as shown by the 41 studies that met our broad inclusion criteria in this scoping review (Figure 2). Studies reporting the co-occurrence of phenotypes with UCL/P sidedness spanned four decades, whereas studies focusing specifically on the cause, or outcomes of UCL/P sidedness were published more recently in the last three decades. In general, included studies were heterogeneous, covering a wide range of study designs, cleft subtypes and outcomes measures and whilst there were trends identified, inconsistencies in the results meant little consensus could be reached. We did not perform a quality assessment of the included studies but observe that more than half of the study designs (n=21) were descriptive in nature (case studies, case series, and cross-sectional studies) with small sample sizes and for the most part without the adjustment for potential confounding factors.

**Figure 2:**
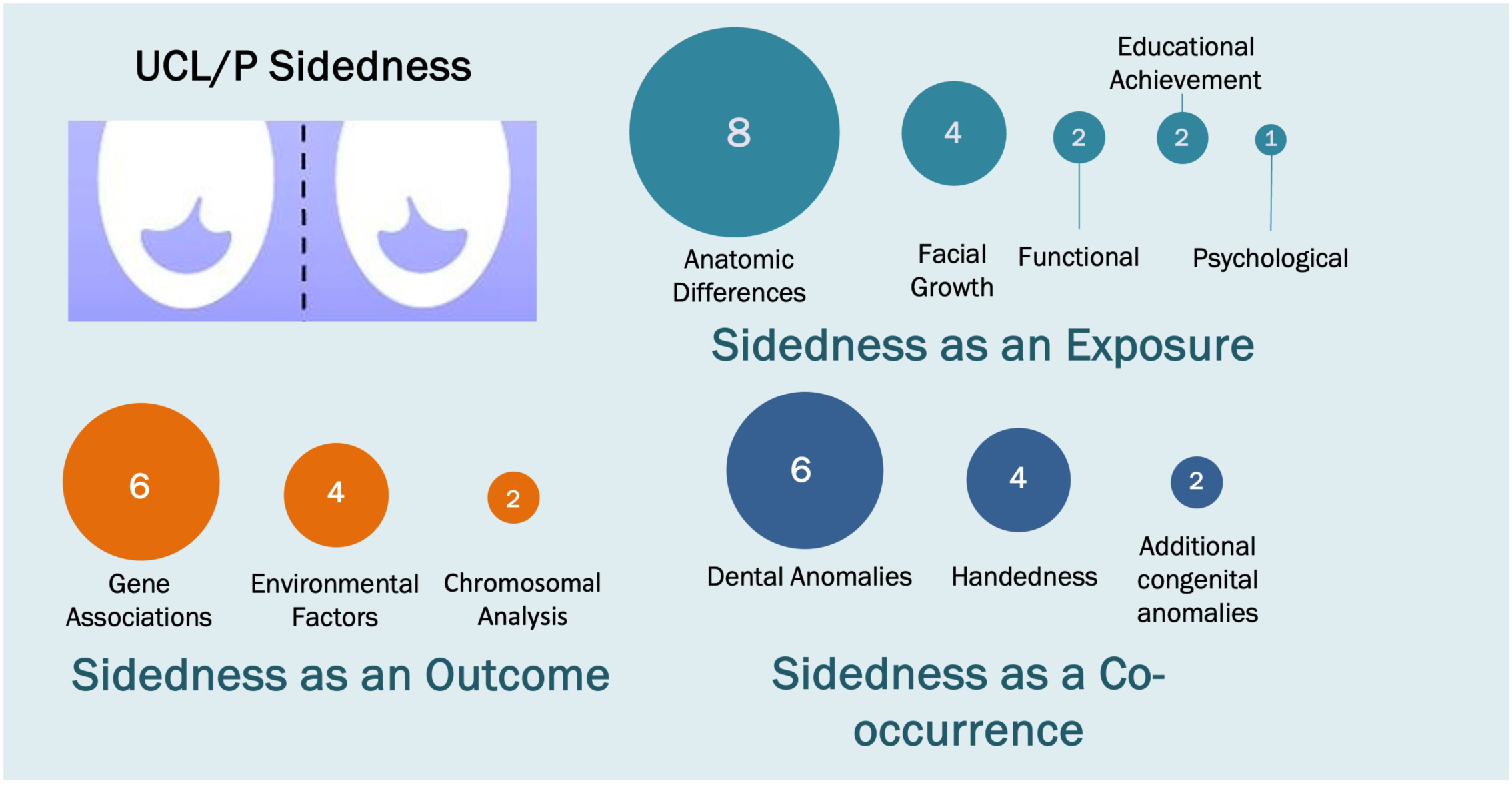
A mapping diagram to show the variables reported relating to sidedness in unilateral cleft lip with or without cleft palate.

Studies reporting left and right UCLP co-occurring with other phenotypes investigated congenital dental anomalies, handedness and additional congenital anomalies. The single area of undisputed consensus was a higher prevalence of additional congenital anomalies in right sided UCLP reported in two independent studies and populations ^27,28^.

Studies investigating cleft sidedness as an outcome reported genetic and environmental exposures associated with either left or right sided UCL/P. These studies suggest that distinct etiologies are possible for left versus right sided UCL/P and furthermore are likely different for sidedness in UCL+-A versus UCLP.

Studies investigating left and right sided UCLP as an exposure covered a variety of outcome measurements. Despite the difficulty of identifying trends due to the heterogeneity of outcomes reported, most studies reported either worse outcomes in right sided UCL/P or no difference in outcomes by UCL/P sidedness. A single study reported a less favorable outcome (educational achievement) for left sided UCL/P ^52^, although this was contradicted by the same first author a year later ^27^.

### Strengths and Limitations

The strengths of this review include a comprehensive systematic search strategy with concerted efforts made to include all languages and a wide variety of study designs. This enabled inclusive mapping of the current literature in UCL/P sidedness. The main limitation of interpreting the results from the scoping review relate to the heterogeneity of the included studies. The broad inclusion of different study designs precluded a quality analysis, although we suspect the overall quality was low.

### Interpretation

Directional asymmetry in UCL/P sidedness is simple and straightforward in terms of its predictable preference for the left side in a 2:1 ratio. In all other respects, cleft sidedness is a fascinating yet complex phenomenon. Our review highlights the promising early findings in this field that has expanded steadily over the past four decades.

In terms of etiology, there was a precedent set in 1942 for cleft subtype differentiation by Fogh-Anderson who demonstrated that cleft palate only is causally different from CL/P ^55^. It is now generally accepted that CL+-A, CLP and cleft palate only have distinct etiologies ^56–58^. Not only have studies within this review suggested it is plausible to consider distinct etiologies for left and right UCL/P but furthermore, the etiology of left versus right sided UCL+-A may be distinct from left versus right sided UCLP ^19,21,28,37^.

Several theories have been postulated about the aetiology of directional asymmetry in UCL/P. The co-occurrence of left and right sided UCL/P with other phenotypes such as congenital dental anomalies, handedness and additional congenital anomalies are of interest because they could potentially have shared or linked aetiologies with UCL/P sidedness. The association between handedness and UCL/P sidedness has been an early focus in the literature because the mesoderm of the face is derived from the neural crest, therefore it is conceivable that there is a connection between cleft aetiology and cerebral hemisphere dominance. Preferential patterns of congenital dental anomalies according to UCL/P sidedness have also drawn interest in the literature because the development of tooth germs and the occurrence of cleft have close embryological relationships in terms of timing and anatomical position ^19^, but although patterns have been found, little conclusive evidence has been added to our overall understanding of laterality causality. The association with other congenital anomalies is of interest because lip formation occurs embryologically at the same time as organogenesis ^7^. The consistent finding of right sided UCLP being associated with a higher prevalence of congenital anomalies in two studies suggests distinct embryological events between the formation of right and left orofacial clefts involving both the primary and secondary palate ^27,28^. Whilst asynchronous embryological fusions have indeed been described mainly in mouse models of embryological palate formation, it does not explain the consistent left sided preference ^59^.

Proposed aetiological mechanisms involved in UCL/P sidedness have included specific genetic and environmental factors, hypoxia and the multifactorial threshold model. Discordant laterality of UCL/P in monogynous twins suggests that perhaps there is differential gene expression in the setting of an identical genotype ^8^. Differential gene expression has been shown to play an important role in the lateralization of the body during embryology ^4^ and studies included in this review have suggested that the FAT4 and FGFR3 genes may be involved in cleft sidedness, but studies have been underpowered to conduct a full GWAS of left versus right sided UCL/P. The variety of genes and chromosomes implicated for both left and right UCL/P suggest a complex polygenic phenomenon, which fits with our current understanding of cleft aetiology. Further highly powered genetic studies are required to add clarity in this area.

Environmental factors such as cigarette smoking may have an impact on cleft aetiology through mechanisms such as hypoxia ^59,60^, which has been shown in animal models to render one side more susceptible to the development of an orofacial cleft ^17^. However, the inconsistencies between the associations of environmental factors to either left or right sided UCL/P makes any conclusions here challenging.

The Multifactorial Threshold Model (MFTM) has been used to describe inheritance in orofacial cleft since the 1990s and involves numerous genes interacting in a multiplicative manner giving rise to an orofacial cleft occurring when the threshold is reached. The MFTM accepts a complex interplay of genetic and environmental factors but argues for a variable proportion of influence depending on the cleft subgroup. According to the model, right sided UCL/P are the less common trait and may represent a more severe phenotype, requiring a higher risk threshold and greater number of genetic and environmental factors ^52^. Right sided UCL/P has been suggested to be on a spectrum of phenotypic severity between a left sided UCL/P and a bilateral CL/P ^22,59^. The consistent finding of a higher prevalence of additional congenital anomalies in right sided UCLP in two studies in this review may lend support to the MFTM ^27,28^. The MFTM has been challenged in the literature ^61^ and indeed it does not help to explain why the prevalence of additional congenital anomalies is higher in RCLP compared to RCL+-A ^28^. Overall, evidence from this review cannot substantiate, nor definitively refute any of the current theories or mechanisms proposed for the aetiology of UCL/P sidedness.

In terms of outcomes impacted by UCL/P sidedness, our knowledge is limited by the relatively few publications in this field and the heterogeneity of outcomes studied. For anatomical differences in particular, a wide array of measurements were reported with only a minority showing differences between left and right sided UCL/P. Perhaps the subtle differences in outcomes between left and right sided UCL/P have failed to attract clinical research. Alternatively, the length of time needed to fully appreciate outcomes in cleft care may mean that few studies looking at long term outcomes are large enough to investigate this. Further work can now be focused on the measures that have been found to be different to help substantiate them.

We report a trend in results reported for either worse outcomes in right sided UCL/P or equivalent outcomes by UCL/P sidedness, with the proviso that a quality analysis was not performed. Anatomical and growth studies reporting worse outcomes for right sided UCL/P have reported a greater degree of hypoplasia in multiple tissue types compared to left sided UCL/P ^41,47,48,50^. Going forward it may help to advance our knowledge of outcomes in patients with UCL/P if awareness is raised amongst the community that sidedness in UCL/P could have a greater impact than purely in the administrative nomenclature of cleft classification.

### Summary and Recommendations

To summarize our findings according to our three initial objectives: first, this scoping review demonstrates an expanding evidence base in the field of sidedness in UCL/P, although there are still only a small number of studies investigating this. Second, studies focusing on aetiology have highlighted many examples to demonstrate different causes of left versus right UCL/P, although pathways remain poorly defined and understood. Third, there are trends for different outcomes when stratified by UCL/P sidedness, although these are currently inconsistent.

We make the following recommendations:

1. Awareness should be raised amongst cleft clinicians and researchers that there could be distinct etiologies and outcomes between left and right sided UCL/P. To that effect, left and right UCL/P should be formally considered as separate cleft subtypes in both clinical and research settings.
2. Furthermore, there is a suggestion that the cause (and therefore potentially the outcome) of UCL/P sidedness may be distinct between UCL+-A and UCLP. Sidedness should ideally be considered separately in research settings for UCL+-A and UCLP, as opposed to grouping together as UCL/P.
3. Sufficiently powered studies are required, because of both the reduced incidence of right sided UCL/P and to differentiate the potentially subtle differences between left and right sided UCL+-A and UCLP.
4. Investment is required in this field to broaden the omics methodologies used. Methodologies demonstrating prior success in differentiating cleft subtypes, such as DNA methylation and large scale GWAS studies could be illuminating.
5. Studies should be explicit about whether they include patients with additional congenital anomalies as the exclusion of patients is likely to skew the results of studies investigating UCL/P sidedness.
6. Confounding factors should be built into the analytic models to reduce the risk of bias. Sex should be given a high priority as a confounding factor in sidedness research as cleft subtypes are known to differ by sex, with UCLP being twice as common in males and cleft palate only being twice as common in females ^62^. Sex has been shown to influence the results as a co-variable in a laterality study included in this review ^60^.

## Data Availability

All data produced in the present work are contained in the manuscript

